# Effectiveness of the BNT162b2 (Pfizer-BioNTech) and the ChAdOx1 nCoV-19 (Oxford-AstraZeneca) vaccines for reducing susceptibility to infection with the Delta variant (B.1.617.2) of SARS-CoV-2

**DOI:** 10.1101/2021.10.12.21264840

**Authors:** Karan Pattni, Daniel Hungerford, Sarah Adams, Iain Buchan, Christopher P Cheyne, Marta García-Fiñana, Ian Hall, David M Hughes, Christopher Overton, Xingna Zhang, Kieran J. Sharkey

## Abstract

**Background:** From January to May 2021 the alpha variant (B.1.1.7) of SARS-CoV-2 was the most commonly detected variant in the UK, but since then the Delta variant (B.1.617.2), first detected in India, has become the predominant variant. The UK COVID-19 vaccination programme started on 8th December 2020. Most vaccine effectiveness studies to date have focused on the alpha variant. We therefore aimed to estimate the effectiveness of the BNT162b2 (Pfizer-BioNTech) and the ChAdOx1 nCoV-19 (Oxford-AstraZeneca) vaccines in preventing infection with respect to the Delta variant in a UK setting.

**Methods:** We used anonymised public health record data linked to infection data (PCR) using the Combined Intelligence for Population Health Action resource. We then constructed an SIR epidemic model to explain SARS-CoV-2 infection data across the Cheshire and Merseyside region of the UK.

**Results:** We determined that the effectiveness of the Oxford-AstraZeneca vaccine in reducing susceptibility to infection is 39% (95% credible interval [34,43]) and 64% (95% credible interval [61,67]) for a single dose and a double dose respectively. For the Pfizer-BioNTech vaccine, the effectiveness is 20% (95% credible interval [10,28]) and 84% (95% credible interval [82,86]) for a single-dose and a double dose respectively.

**Conclusion:** Vaccine effectiveness for reducing susceptibility to SARS-CoV-2 infection shows noticeable improvement after receiving two doses of either vaccine. Findings also suggest that a full course of the Pfizer-BioNTech provides the optimal protection against infection with the Delta variant. This would advocate for completing the full course programme to maximise individual protection and reduce transmission.

## 1 Background

The UK COVID-19 vaccination programme started on the 8th December 2020 and, by 19 September 2021, the overall vaccine uptake for 1 dose was 89.3% and 83.9% for 2 doses in England for adults aged 18 and over [1]. Assessing the effectiveness of the vaccines is important for government policy, and particularly so as more transmissible variants of SARS-CoV-2 emerge [2]. The current dominant SARS-CoV-2 variant in the UK is Delta (B.1.617.2) [3], and more recent vaccine effectiveness studies now focus on this variant [4, 5, 6].

Direct vaccine effectiveness is often estimated using a test-negative case-control design, which compares the odds of vaccination in a group of symptomatic individuals that test positive for COVID-19 with the control group who are defined as individuals showing symptoms of COVID-19 but test negative. This methodology was employed in two recent COVID-19 vaccine effectiveness studies conducted in England and Scotland [4, 6]. A study by Pouwels et al., 2021 used a more traditional case-control design with survey data from randomly selected households across the UK [5]. Here, the control group consisted of randomly selected individuals who did not contract COVID-19. Test-negative designs are often logistically beneficial and cost-effective and can help to minimise selection bias because the cases and controls are assumed to have similar health-seeking behaviour. But, one of the issues raised with test-negative case-control designs is the lack of generalisability [7, 8]; that is, it only considers individuals who have sought to get tested and, therefore, findings may not be generalisable to those individuals who did not access testing services.

Vaccine effectiveness can also be estimated using a compartmental epidemic model that accounts for vaccination. Various methods of forecasting SARS-COV-2, including compartmental models, have thus far have been used to study hypothetical scenarios [9]. For example, Wong et al., 2021 used the SIR (susceptible, infected or removed/recovered) model to consider a single dose vaccination program and used it to project new SARS-CoV-2 cases based on different vaccine effectiveness levels [10]. Another example is the modified version of the SEIR (where E represents a compartment for exposed individuals) model with a single dose vaccine program, and waning natural and vaccine-induced immunity, which is used to study different vaccination policies [11].

Here we consider a modified SIR model with a multi-dose vaccine program to estimate vaccine effectiveness for the BNT162b2 (hereafter referred to as Pfizer-BioNTech) and ChAdOx1 nCoV-19 (hereafter referred to as Oxford-AstraZeneca) vaccines for reducing susceptibility to infection with respect to the Delta variant in England, UK. The method we consider here is not restricted to those individuals who have been tested but considers the entire resident population, therefore providing a more generalisable population estimate for vaccine effectiveness. The method also explicitly accounts for the temporal variation in vaccination levels as well as levels of infection in the population, removing these as a potential source of bias. We exploit a specific time window where initially low levels of infection are being driven rapidly upwards by the emergence of the Delta variant, justifying the use of a simple SIR model.

## 2 Methods

### 2.1 Data

We used data from the Combined Intelligence for Population Health Action (CIPHA; www.cipha.nhs.uk) data resource. CIPHA covers the population health management of ∼ 2.6M General Practice registered population of Cheshire and Merseyside, UK. It includes person-level linked anonymised records across the National Health Service (NHS), local government, social care, administrative and public health information systems. From CIPHA we have detailed case data for SARS-CoV-2 PCR positive individuals together with individual-level vaccination data. For demographics of data see Additional file 1: Table S1.1.

Demographic data and our denominator population was for the whole of Cheshire and Merseyside and was taken from the general practice registered population, sourced from the Spine Demographics service. SARS-CoV-2 PCR testing data came from the Public Health England (PHE) Second Generation Surveillance System (SGSS) feeds. For this work this consisted of all Pillar 1 (swab testing in PHE labs and NHS hospitals) and Pillar 2 (swab testing for the wider population, as set out in government guidance) tests taken by individuals whose home address was registered within Cheshire and Merseyside [12]. We considered only SARS-CoV-2 PCR positive cases in this study and vaccination status data came from the National Immunisation Management System (NIMS). All of these data feeds came via the CIPHA platform.

### 2.2 SIR model with vaccination

We use an SIR model [13, 14] where individuals are also classified according to their vaccination status. We do not consider an age stratified model to keep parameters to a manageable level. There is some age effect in vaccine distribution (Figure 2) but we consider this sufficiently small for this simplification to be used. Susceptible, infected and removed individuals are respectively denoted *S, I*, and *R* when unvaccinated, and *S*_*ij*_, *I*_*ij*_, and *R*_*ij*_ when vaccinated, where *i* is the number of doses and *j* the type of vaccine. Considering a two-dose vaccine program, i.e. *i* ∈ {1, 2}, the flows between the various classes are shown in Figure 1 and the system of differential equations is given by:

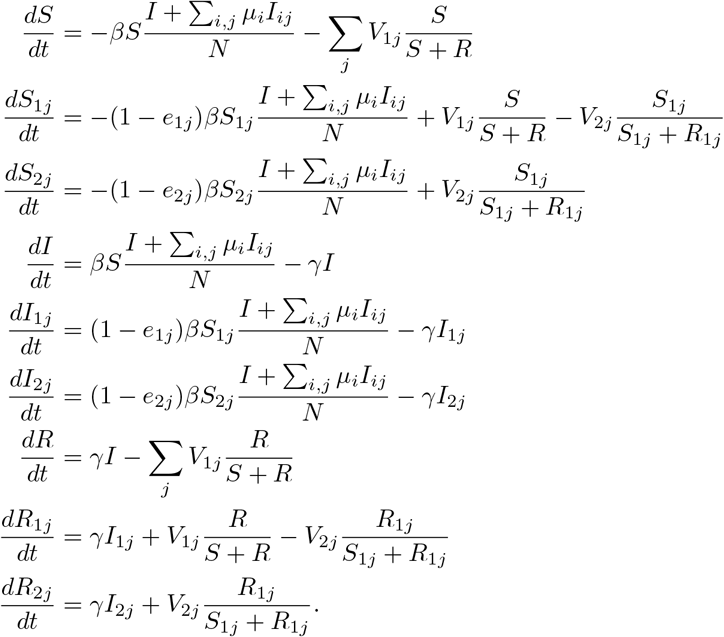

**Figure 1:**
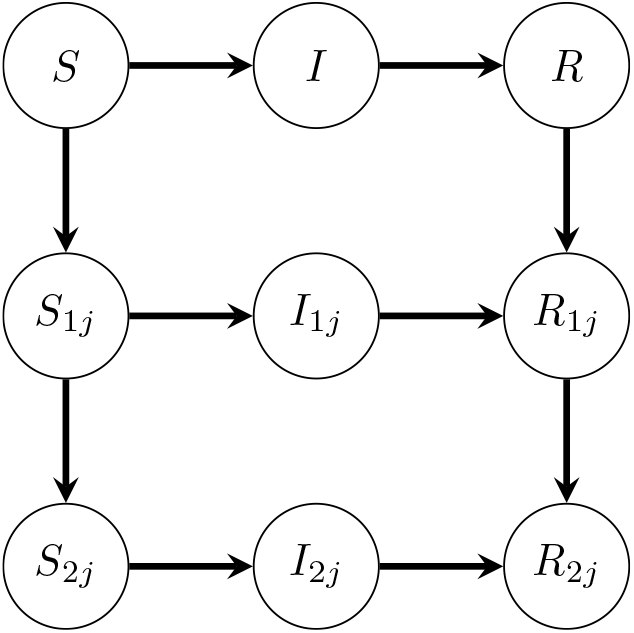
The transitions between the various classes of individuals, namely susceptible (*S* and *S*_*ij*_), infected (*I* and *I*_*ij*_) and removed (*R* and *R*_*ij*_) when considering a two-dose vaccine program. *S, I, R* are unvaccinated and *S*_*ij*_, *I*_*ij*_, *R*_*ij*_ are vaccinated where *i* is the number of doses received and *j* is the type of vaccine.

**Figure 2:**
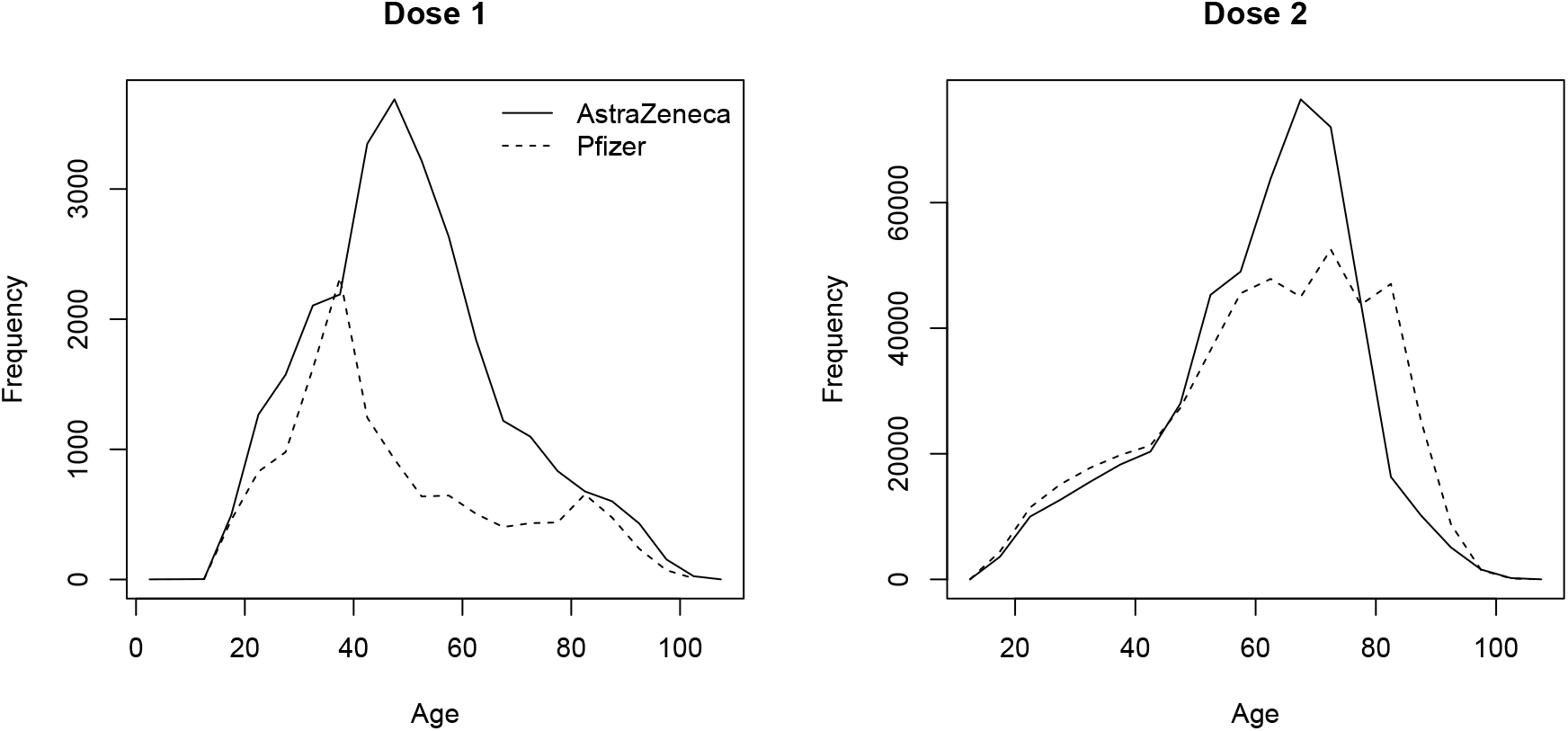
Histogram showing age distribution for Pfizer-BioNTech and Oxford-AstraZeneca vaccines on and before 24 May 2021.

As shown in Figure 1, the flow of individuals in this model is not only from susceptible to infected to removed, but also unvaccinated to one-dose to two-doses within the susceptible and removed classes. Birth and death processes are neglected in this model. The transmission rate, *β*, is the rate at which an unvaccinated infected individual (*I*) transmits the virus to an unvaccinated susceptible individual (*S*). For vaccinated infected individuals (*I*_*ij*_), their infectiveness, i.e. how likely they are to infect a susceptible individual, is assumed to be reduced by a factor *µ*_*i*_ after *i* doses and we make no distinction between the vaccine types; the conclusions presented here are shown to be very insensitive to this parameter and hence to differences in this parameter between vaccine types. In vaccinated susceptible individuals (*S*_*ij*_), the effectiveness of dose *i* of vaccine *j* in preventing infection is *e*_*ij*_. The recovery rate, *γ*, of an infected individual is assumed to be the same regardless of their vaccination status. As can be seen from the form of the equations for infectious individuals, we expect this parameter to be highly correlated with *β* and so not well-constrained by the data. We investigate sensitivity to this parameter over a wide range of plausible values and show that our conclusions on vaccine effectiveness are not sensitive to this. The quantity *V*_*ij*_ is the rate of vaccination with dose *i* of vaccine *j* and is determined from the CIPHA data to give a daily vaccination rate. We assume that *V*_1*j*_ is evenly distributed to individuals in classes *S* and *R, V*_2*j*_ is evenly distributed to the individuals in classes *S*_1*j*_ and *R*_1*j*_, and that infected individuals do not receive the vaccine. The basic SIR model is recovered by initialising all vaccinated populations at zero and setting *V*_*ij*_ = 0 for all *i, j*. A summary of the notation used is given in Table 1.

**Table 1:** Table of Notation.

| Notation | Definition |
| --- | --- |
| $\beta$ | Transmission Rate. |
| $\gamma$ | Rate of recovery ($\text{days}^{-1}$). |
| $e_{ij}$ | Percentage effectiveness against infection of $i$ doses of vaccine $j$. |
| $V_{ij}$ | Daily number of individuals who have received dose $i$ of vaccine $j$. |
| $N$ | Number of individuals in the population. |
| $\mu_i$ | Factor reducing infectiveness due to $i$ doses. |

For simplicity, we do not include an exposed state in our model since this would increase the number of fit parameters, lose information and increase instability. Adding a short delay between infection and infectiousness is unlikely to impact the parameters of interest which only concern the rate of exponential growth of the infectious population and previous analysis has shown that the type of data we investigate is more reliably analysed using an SIR model rather than an SEIR model [15].

### 2.3 Model fitting

On 17th May 2021, indoor hospitality was reopened in England. There was a spike in the number of covid-19 cases due to the dominance of the Delta variant in the Cheshire and Merseyside NHS region (see Figure 3 (a)) and shown elsewhere [3] in combination with the lifting of restrictions. To construct Figure 3 (a), a cycle threshold (Ct) cut-off of *≤* 35 [16] for all 3 genes (see [17] for details) required to determine whether it is the Delta or another variant. Only data from processing labs that routinely looked at the 3 genes was used. At this point in time, 49% of the population had received 1 dose and 24% had received 2 doses in this region (see Figure 3 (b)). We use the rapid growth in infections during the period following this date to estimate the effectiveness of the Oxford-AstraZeneca and Pfizer-BioNTech vaccines in our model. During the period of our analysis, use of other vaccines was negligible (of the total vaccines administered, less than 2% of dose 1 and less than 1% of dose 2 were the Moderna vaccine in the CIPHA dataset and no other vaccine types were used) and there were no instances of individuals receiving two different vaccines.

**Figure 3:**
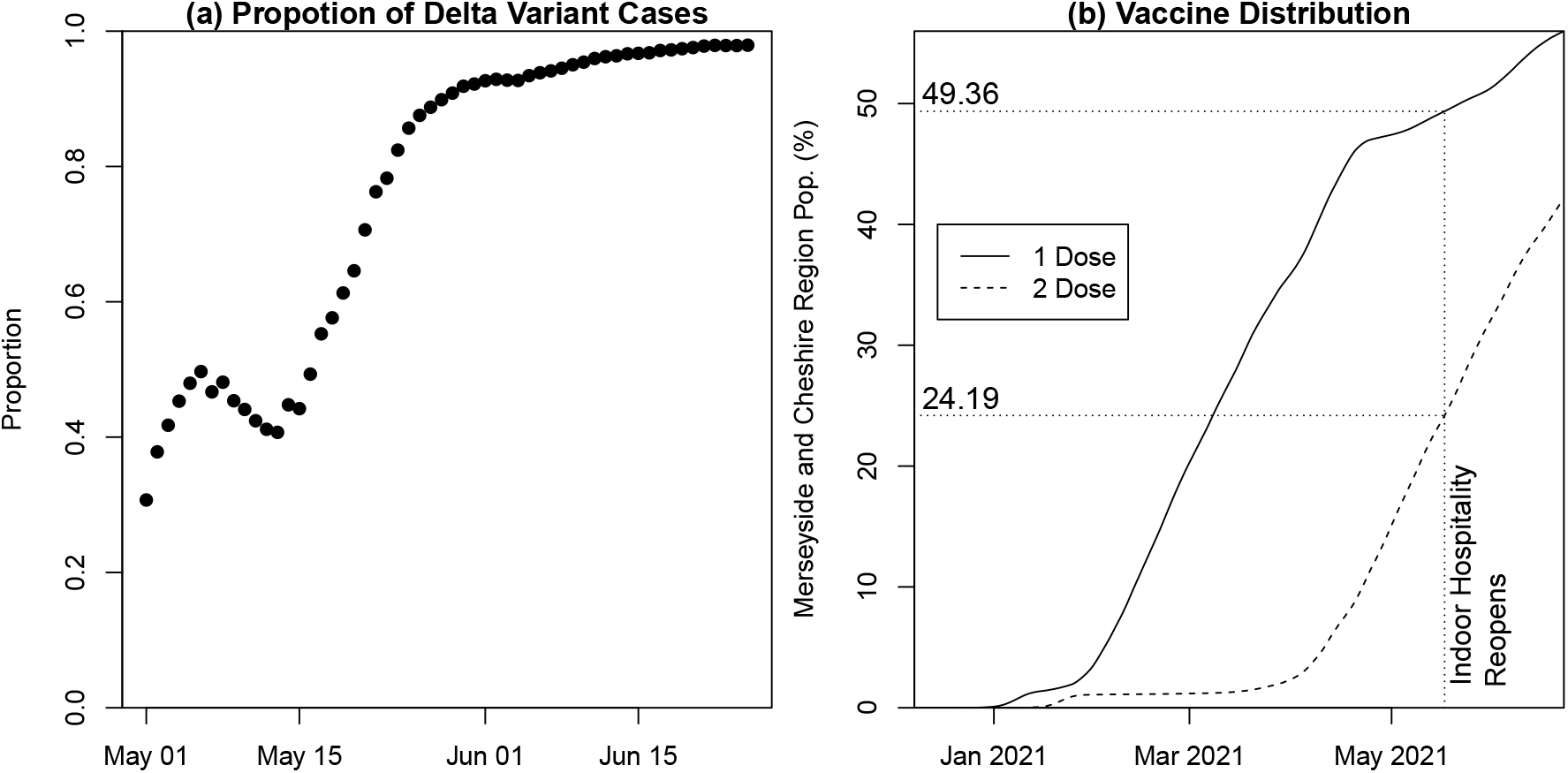
(a) Proportion of Delta variant cases between 1st May 2021 and 25th June 2021. (b) Vaccine distribution in Cheshire and Merseyside region.

The fitting window used is shown in Figure 4 (a). The fitting window starts on 24th May 2021, 7 days after indoor hospitality was reopened. This accounts for the delay in symptoms emerging, which is when people are likely to get tested for COVID-19 [18, 19] and also accounts for the 7 day symmetric rolling average, where the number of cases on a given day, 3 days before and 3 days after are averaged. We do this to smooth out the pronounced variations in reporting rates over the course of a week. We can have a symmetric rolling average since this is historic data but note that this differs from rolling averages computed for current data which necessarily involves the 6 preceding days [20].

**Figure 4:**
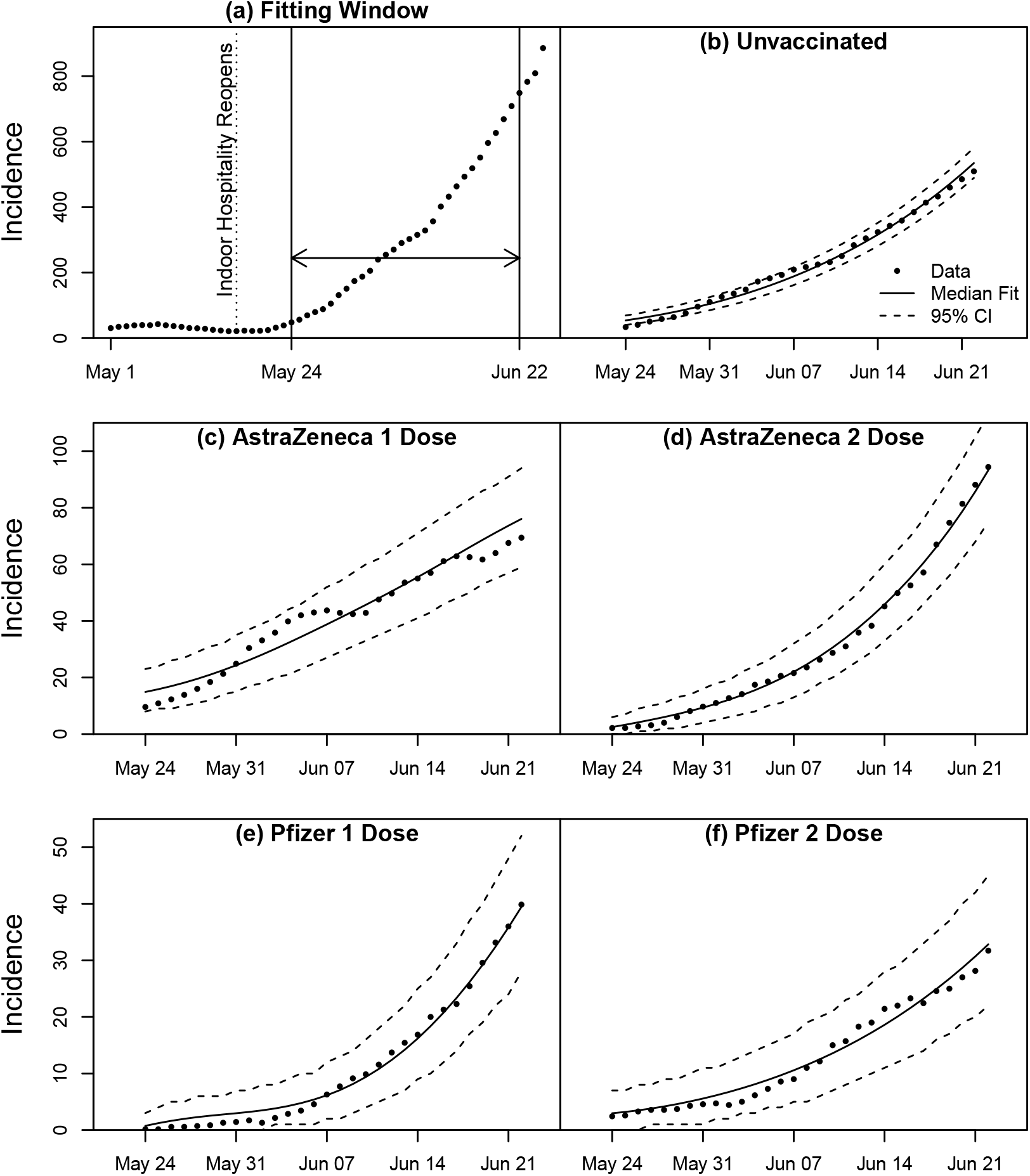
(a) Fitting window for 7-day rolling averaged data for all incidence. (b)-(f) SIR model with multi-dose vaccine and median parameter values from MCMC fitting analysis overlayed on 7-day rolling averaged incidence data for different vaccinations states.

Markov Chain Monte Carlo (MCMC) methods were used to fit to the incidence and vaccination time series data using the R-statistical package BayesianTools [21]. The values used to initialise the model fits are shown in Table 2. For parameters that are estimated by model fitting, a value was randomly chosen from their prior distribution to initiate the fits. An adaptive Metropolis-Hastings algorithm was used where the parameter covariance was updated every 500 iterations after a burn-in of 2000 iterations. The algorithm was run for total of 8 *×* 10^5^ iterations excluding burn-in. The final 3 *×* 10^5^ iterations were used to construct the posterior distributions of the parameters, which are plotted in the appendix.

**Table 2:** Prior distributions and initial values (obtained from CIPHA data) used for model fits.

| (a) Prior distributions used for initialising model fits. |  |  |  |  |  |  |  |  |  |
| --- | --- | --- | --- | --- | --- | --- | --- | --- | --- |
| Name | $\gamma$ | Prior Distributions | | Initial Values | | | | | |
| | | $\beta, I, I_{ij}$ | $e_{ij}, \mu_i$ | $S, S_{ij}, R, R_{ij}$ | | | | | |
| Core Model | 1/7 | Unif[0, ∞] | Unif[0, 1] | Table 2 (b) | | | | | |
| Sensitivity to $\gamma$ | 1/3 | Unif[0, ∞] | Unif[0, 1] | Table 2 (b) | | | | | |
| Sensitivity to $\gamma$ | 1/11 | Unif[0, ∞] | Unif[0, 1] | Table 2 (b) | | | | | |
| Double Removed | 1/7 | Unif[0, ∞] | Unif[0, 1] | Table 2 (c) | | | | | |

| (b) Initial values when assuming all infected from start of pandemic moved to removed class. |  |  |  |  |  |  |  |  |  |
| --- | --- | --- | --- | --- | --- | --- | --- | --- | --- |
| $S$ | $S_{1A}$ | $S_{2A}$ | $S_{1P}$ | $S_{2P}$ | $R$ | $R_{1A}$ | $R_{2A}$ | $R_{1P}$ | $R_{2P}$ |
| 1266979 | 478347 | 312594 | 74665 | 384481 | 70887 | 42715 | 20895 | 6738 | 26715 |

| (c) Initial values when assuming infected individuals are underestimated by 50%. |  |  |  |  |  |  |  |  |  |
| --- | --- | --- | --- | --- | --- | --- | --- | --- | --- |
| $S$ | $S_{1A}$ | $S_{2A}$ | $S_{1P}$ | $S_{2P}$ | $R$ | $R_{1A}$ | $R_{2A}$ | $R_{1P}$ | $R_{2P}$ |
| 1196092 | 435632 | 291699 | 67927 | 357766 | 141774 | 85430 | 41790 | 13476 | 53430 |

$\beta$ : Transmission Rate, $\gamma$ : Recover Rate, $e_{ij}$ : Effectiveness of $i$ doses of vaccine $j$ , $S$ : Unvaccinated susceptible individuals, $S_{ij}$ : Susceptible individuals who have received $i$ doses of vaccine $j$ , $I$ : Unvaccinated infected individuals, $I_{ij}$ : Infected individuals who have received $i$ doses of vaccine $j$ , $R$ : Unvaccinated removed individuals, $R_{ij}$ : Removed individuals who have received $i$ doses of vaccine $j$ , $\mu_i$ : Infectiveness of an infected individual with $i$ doses of either vaccine. For number of doses we have $i \in \{1, 2\}$ and for vaccines $j \in \{A, P\}$ for Oxford-AstraZeneca and Pfizer-BioNTech respectively.

The likelihood function used for MCMC fitting is the Negative Binomial function as in [22]. The Negative Binomial probability mass function is

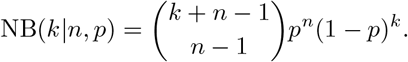

The following parameterisation is used:

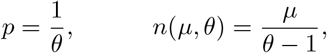

where *µ* is the mean of the distribution and the variance is *µθ*. Let **I**(*t*) = [*I*(*t*), *I*_*ij*_(*t*)] for all *i, j* be the observed daily incidence on day *t* and **Î**(*t, x*) be the incidence generated by the model on day *t* for a given set of model parameters *x*. It is assumed that

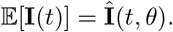

The log likelihood function is then given by

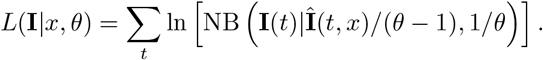

The SIR model with multi-dose vaccines is fitted to the data to estimate the posterior distributions of the transmission rate (*β*), the effectiveness of the vaccines (*e*_*ij*_), the initial values of the infected classes (*I* and *I*_*ij*_), and infectiousness (*µ*_*i*_). We have *i* ∈ {1, 2} and *j* ∈ {*A, P*} for Oxford-AstraZeneca and Pfizer-BioNTech respectively, and therefore posterior distributions for 12 parameters are estimated by the model fit. For all model fits we use uniformly distributed priors for all 12 parameters estimated as shown in Table 2(a). For the impact of vaccines on infectiousness (*µ*_*i*_), which implements the reduction in the ability of an individual to transmit the virus, we have no data for the Delta variant of COVID-19. For the Alpha variant, this has been estimated to be 0.45-0.50 for one dose of Pfizer-BioNTech and 0.35-0.50 for one dose of Oxford-AstraZeneca, with no data available for the second dose [23]. We therefore make no *a priori* assumption about infectiousness and consider its full range for the prior distribution; i.e. Unif[0, 1], allowing for analysis of sensitivity to these parameters. For vaccine effectiveness (*e*_*ij*_), we also consider the full viable range for the prior distribution; i.e. Unif[0, 1]. The transmission rate (*β*), initial *I* and initial *I*_*ij*_ cannot be negative so a lower bound of 0 is used for the prior distribution, i.e. Unif[0, *∞*].

The infectious period in days is given by 1/*γ*, with estimates ranging between 3 and 11 days according to [24]. We therefore consider three model fits with regards to *γ* (see Table 2 (a)). For our ‘Core Model’ fit we use a median infectious period of 7 days, i.e. *γ* = 1/7. To account for sensitivity to the infectious period, we also fit the model for *γ* = 1/3 and *γ* = 1/11; these fits are called ‘Sensitivity to *γ*’. For these three fits, the fixed parameters (*V*_*ij*_, *N*) and the initial values of *S, S*_*ij*_, *R, R*_*ij*_, are obtained from CIPHA data. For *V*_*ij*_ we take into account a lag for the vaccine to come into effect. An individual is assumed to move into the relevant vaccinated category after a delay of 21 days post-vaccination for dose 1 and 14 days post-vaccination for dose 2 of either vaccine [25]. The population size for our data covering Cheshire and Merseyside on 24th May 2021 is *N* = 2 691 418. This is taken from the CIPHA database where we remove deceased individuals from any cause. We assume this number is fixed for the duration of the fit window. The initial values *S*_*ij*_ and *R*_*ij*_ are shown in Table 2 (b). The number removed are taken from all individuals who have ever been recorded as infected since the beginning of the pandemic, except for those that died. Whilst infection-blocking immunity wanes rapidly, disease-reducing immunity is long-lived [26]. We are therefore assuming immunity for the remaining duration of the pandemic, and we are assuming that there is no under-reporting of cases (which is certainly not true, especially during the first wave of the pandemic) [27]. The first of these assumptions leads to an overestimate of the removed category on 24th May 2021, and the second (likely more questionable) assumption leads to an underestimate of the removed category. We demonstrate insensitivity of our conclusions to these assumptions by re-running our analysis assuming that only half of all the infected were actually detected overall, leading to a doubling of the initial removed category on 24th May 2021. This fit is called ‘Double Removed’ (see Table 2 (c)).

## 3 Results

Our model used a total population of 2,730,111 (Additional File Table S1.1). From the Core Model, the effectiveness obtained for one dose is 38.5% (95% credible interval [34.3,42.6]) for Oxford-AstraZeneca and 19.5% (95% credible interval [10.4,28.1]) for Pfizer-BioNTech. For two doses, we obtained an effectiveness of 64.0% (95% credible interval [61.4,66.5]) for Oxford-AstraZeneca and 83.9% (95% credible interval [82.1,85.6]) for Pfizer-BioNTech. The median value and and 95% credible interval for all fitted parameters are shown in Table 3 for the Core Model, and Additional File 1: Table S2.1 for all model fits. The MCMC trace plots and posterior distributions for all model fits are shown in the Additional File 2: Figures S2.1–S2.4.

**Table 3:** Estimates obtained from Core Model (*γ* = 1/7).

| Parameter | Median (50%) | Lower (2.5%) | Upper (97.5%) |
| --- | --- | --- | --- |
| $\beta$ | 0.4060 | 0.3893 | 0.4425 |
| $e_{1A}$ | 0.3851 | 0.3425 | 0.4260 |
| $e_{2A}$ | 0.6402 | 0.6140 | 0.6647 |
| $e_{1P}$ | 0.1954 | 0.1041 | 0.2809 |
| $e_{2P}$ | 0.8392 | 0.8212 | 0.8559 |
| Initial $I$ | 54.0151 | 48.4328 | 59.5435 |
| Initial $I_{1A}$ | 14.8932 | 10.3012 | 19.8382 |
| Initial $I_{2A}$ | 2.4775 | 0.9300 | 4.7359 |
| Initial $I_{1P}$ | 0.7469 | 0.1234 | 2.0681 |
| Initial $I_{2P}$ | 2.9717 | 1.3856 | 5.1267 |
| $\mu_1$ | 0.8836 | 0.4464 | 0.9955 |
| $\mu_2$ | 0.0545 | 0.0021 | 0.2734 |

$\beta$ : Transmission Rate, $e_{ij}$ : Effectiveness of $i$ doses of vaccine $j$ , Initial $I$ : Initial number of unvaccinated infected individuals, Initial $I_{ij}$ : Initial number of infected individuals who have received $i$ doses of vaccine $j$ , $\mu_i$ : Infectiveness of an infected individual with $i$ doses of either vaccine. For number of doses we have $i \in \{1, 2\}$ and for vaccines $j \in \{A, P\}$ for Oxford-AstraZeneca and Pfizer-BioNTech respectively.

Figure 4 (b)–(f) shows the Core Model fit to data. The incidence curves and 95% confidence interval (CI) bands are plotted using the Core Model median parameter values (Table 3) together with the incidence curves given by the data. The 95% CI is generated by using the fact that the likelihood function used for MCMC fitting is the Negative Binomial function.

For all model fits (see Table 2 (a)), the parameters of direct interest (the vaccine effectiveness parameters) are reproduced in Table 4 where other results from existing studies are reproduced for direct comparison. For our results we have stated the median value together with the 95% credible interval, where as for the results in [4, 5, 6] the mean value together with the 95% confidence interval is given.

**Table 4:**
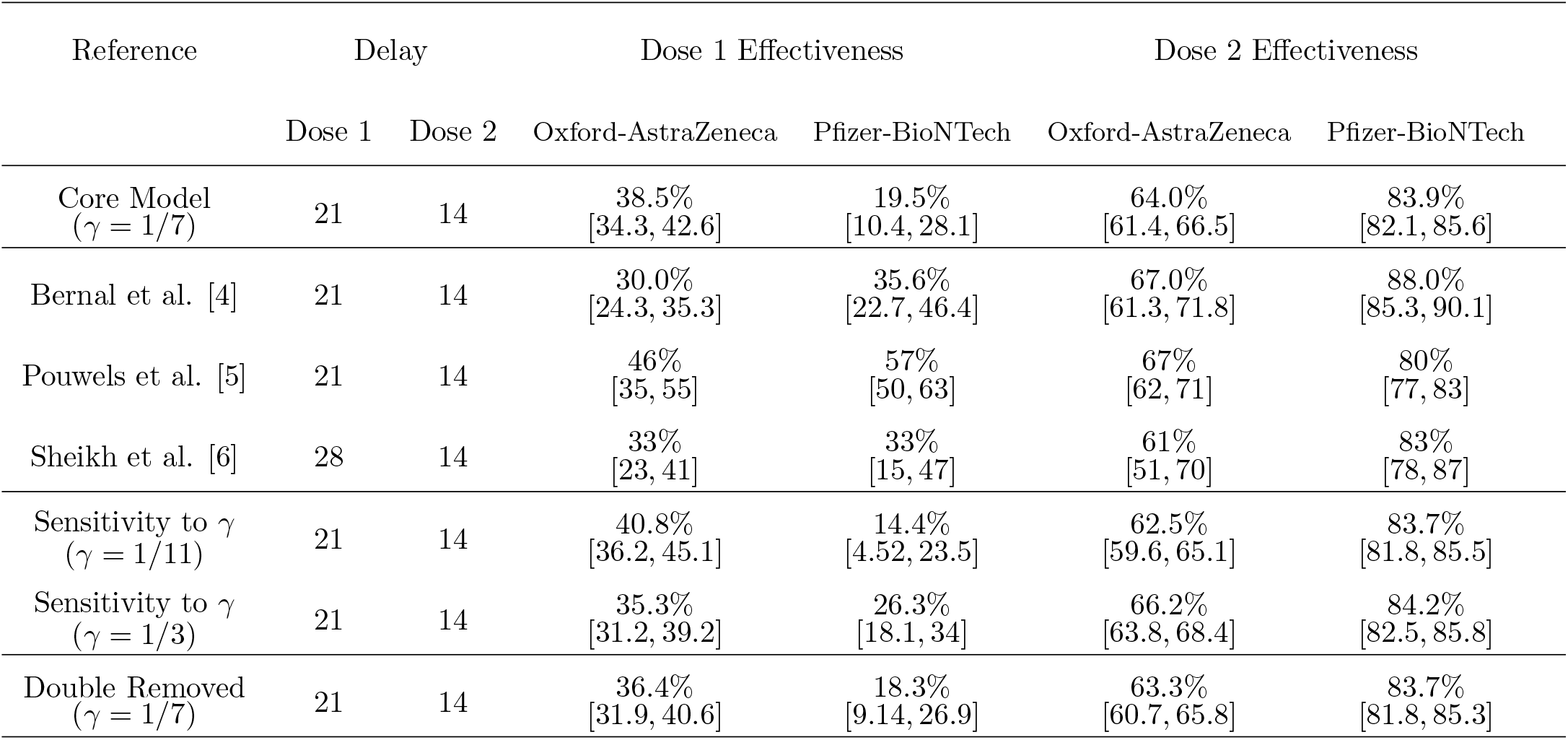
Vaccine effectiveness against Delta variant (Delay is the number of days after which the effectiveness is measured, and *γ* is the recovery rate.).

| Reference | Delay |  | Dose 1 Effectiveness |  | Dose 2 Effectiveness |  |
| --- | --- | --- | --- | --- | --- | --- |
|  | Dose 1 | Dose 2 | Oxford-AstraZeneca | Pfizer-BioNTech | Oxford-AstraZeneca | Pfizer-BioNTech |
| Core Model<br>($\gamma = 1/7$) | 21 | 14 | 38.5%<br>[34.3, 42.6] | 19.5%<br>[10.4, 28.1] | 64.0%<br>[61.4, 66.5] | 83.9%<br>[82.1, 85.6] |
| Bernal et al. [4] | 21 | 14 | 30.0%<br>[24.3, 35.3] | 35.6%<br>[22.7, 46.4] | 67.0%<br>[61.3, 71.8] | 88.0%<br>[85.3, 90.1] |
| Pouwels et al. [5] | 21 | 14 | 46%<br>[35, 55] | 57%<br>[50, 63] | 67%<br>[62, 71] | 80%<br>[77, 83] |
| Sheikh et al. [6] | 28 | 14 | 33%<br>[23, 41] | 33%<br>[15, 47] | 61%<br>[51, 70] | 83%<br>[78, 87] |
| Sensitivity to $\gamma$<br>($\gamma = 1/11$) | 21 | 14 | 40.8%<br>[36.2, 45.1] | 14.4%<br>[4.52, 23.5] | 62.5%<br>[59.6, 65.1] | 83.7%<br>[81.8, 85.5] |
| Sensitivity to $\gamma$<br>($\gamma = 1/3$) | 21 | 14 | 35.3%<br>[31.2, 39.2] | 26.3%<br>[18.1, 34] | 66.2%<br>[63.8, 68.4] | 84.2%<br>[82.5, 85.8] |
| Double Removed<br>($\gamma = 1/7$) | 21 | 14 | 36.4%<br>[31.9, 40.6] | 18.3%<br>[9.14, 26.9] | 63.3%<br>[60.7, 65.8] | 83.7%<br>[81.8, 85.3] |

## 4 Discussion

We assessed the effectiveness of the Oxford-AstraZeneca and Pfizer-BioNTech vaccines in reducing the susceptibility of individuals to the SARS-CoV-2 Delta variant of COVID-19 using data from the Cheshire and Merseyside NHS region of the UK. We found that both vaccines provide good protection after two doses but substantially less protection after one dose. The one dose effectiveness was greater for Oxford-AstraZeneca (39%) compared to Pfizer-BioNTech (20%), however the Pfizer-BioNTech vaccine provides greater protection against infection with the Delta variant after two doses (84% compared to Oxford-AstraZeneca vaccine 64%). Our estimates of vaccine effectiveness for one dose of Oxford-AstraZeneca and two-doses of either vaccine are consistent with those reported by [4, 5, 6] (Table 4). Even after changing assumptions, which include the infectious period and the number of removed individuals, the results are still consistent.

However, for one dose of the Pfizer-BioNTech vaccine, our estimate is slightly lower than that reported in these studies but still comparable to those reported by Bernal and et al., 2001 and Sheikh et al., 2001 [4, 6]. Similar to our study these studies included cases who had actively sought COVID-19 testing. In contrast, the study conducted by Pouwels et al., 2021 used a community household testing survey to identify cases and controls and notably reported higher 1st dose effect estimates for the Pfizer-BioNTech vaccine [5]. This higher effect estimate may be somewhat explained by age of the population immunised with just one dose of Pfizer-BioNTech vaccine, as they were skewed towards younger individuals compared to our study [5]. This is critical for contextualising the vaccine effect estimates, as a greater immune response and effectiveness have been reported in young populations after one dose of the Pfizer-BioNTech vaccine [28, 29].

Here the effectiveness of the Oxford-AstraZeneca and Pfizer-BioNTech vaccines was estimated by fitting an SIR model where each class of individual was stratified by the number of doses and type of vaccine received. We identified a unique time period in May and June 2021 where the epidemic was undergoing exponential growth from a very low level due to the emergence of the Delta variant and lifting of restrictions, enabling the use of a simple SIR model. During this same period, substantial numbers of vaccines were being administered and this enabled us to extract strong signals for the effectiveness of single and double doses of the Pfizer-BioNTech and Oxford-AstraZeneca vaccines.

The temporal dynamics in each vaccination category are fully accounted for, removing biases caused by the interaction of vaccination rates/ types and the level of infection in the community. For studies employing case-control methodology, these biases are harder to account for, for example, there is an assumption that the vaccine under study has no effect on disease incidence in the control population (i.e. the herd effects) [6]. In addition, our study is not restricted to individuals who have sought to get tested and so minimises the issue of population level generalisability [7, 8].

A major assumption in our model is that the number of removed individuals at the start of the fitting window is given by the actual number of recorded infections throughout the whole pandemic. This could be problematic for two reasons. Firstly, there may be waning immunity of previously infected individuals. Secondly, there may be under-reporting of infections, particularly in the early stages of the pandemic, which could be due to asymptomatic infection, choosing not to get tested, or due to lack of availability of testing. The two effects act in opposite directions and the second is likely to be the most dominant on the present timescale. This means the number of susceptible individuals is likely to be overestimated. To account for this, we considered the case where the number of removed individuals in each class are doubled, representing an under-reporting of 50% [30]. This reduces the number in each respective susceptible class for the initial conditions of our model. We make no similar assumption for the dynamics of infection since current detection rates are likely to be high and the fit parameters of interest (exponential rates) are insensitive to reporting rates provided that these rates are constant during the fit window. The vaccine effectiveness estimates are found to be quite insensitive to even this significant modification.

### 4.1 Limitations

It is important to note that our model does not stratify the population by age and therefore it does not take into account the effects of age on vaccine effectiveness. In the UK, vaccines were initially prioritised for the most vulnerable people and then distributed in decreasing order of age [31]. In the fitting window we have used to estimate vaccine effectiveness, unvaccinated individuals or individuals with one dose are much younger. In particular, there is a greater distribution of one dose amongst those *≤* 50 years, and a greater distribution of 2 doses amongst those *≥* 70 years. This means that the single dose vaccine effectiveness is likely to be biased towards the younger population, whereas those with two doses towards the older population. There may be an effect due to variation in immunity across age groups, where younger individuals are likely to have a better immune response to vaccines. The effectiveness of a given vaccine would therefore also depend upon how it is distributed across different age groups. However, Cheshire and Merseyside has had slower population level COVID-19 vaccine uptake compared to other areas of the UK [1], which has benefits for estimating vaccine effectiveness in post-licensure studies as this has resulted in a more heterogeneous age distribution, particularly in the population which has only received one dose of the Pfizer-BioNTech vaccine.

## 5 Conclusion

Vaccine effectiveness for reducing susceptibility to SARS-CoV-2 infection shows noticeable improvement after receiving two doses of either vaccine. Our findings also suggest that a full course of the Pfizer-BioNTech vaccine provides the optimal protection against infection with the Delta variant. These findings advocate for completion of the full course to maximise individual protection and reduce transmission.

## Data Availability

Pseudonymised data are accessible via Combined Intelligence for Population Health Action (CIPHA). Requests can be made to the Data Asset and Access Group for extracts of the larger-scale data which cannot be released openly due to information governance requirements. All R code is accessible from the corresponding author.

## Acknowledgements

We thank CIPHA for providing access to the data used in this study. KJS and KP acknowledge support from the EPSRC (grant EP/T031727/1). DH is funded by a National Institute for Health Research (NIHR) Post-doctoral Fellowship (PDF-2018-11-ST2-006).

## Abbreviations

AIC: Akaike Information Criterion
CI: Confidence interval
CIPHA: Combined Intelligence for Population Health Action
MCMC: Markov Chain Monte Carlo
NHS: National Health Service
NIMS: National Immunisation Management System
PCR: Polymerase Chain Reaction
RNA: Ribose Nucleic Acid
SARS-CoV-2: Severe acute respiratory syndrome coronavirus 2
SIR: Susceptible, Infectious or Recovered/Removed
SEIR: Susceptible, Exposed, Infectious or Recovered/Removed
UK: United Kingdom

## Author Information

### Contributions

KJS conceptualised and designed the study and was responsible for supervision. KP was responsible for visualisation, formal analysis and writing the original draft. CC, DMH and XZ were responsible for accessing and verifying the data. All authors reviewed and edited the manuscript and read and approved the final manuscript.

## Ethics declarations

### Ethics approval and consent to participate

The University of Liverpool provided secondary data analysis on fully anonymised data. As per the National Health Service (NHS) Health Research Authority guidelines, this work did not require ethical approval. Cheshire & Merseyside Health & Care Partnership Combined Intelligence for Population Health Action (CIPHA) Data Asset and Access Group approved access to the anonomysed data contained in the study. MAST (mass, asymptomatic, serial testing)/SMART (systematic, meaningful, asymptomatic/agile, repeated testing) was defined as ‘an emergency public health intervention during an extraordinary event’ which were subject to the legal and ethical provisions of a health protection activity and COVID-19 specifically. The secondary analysis of data provided in a health protection activity is not classified as research, and so does not require research ethics committee review (see http://www.hra-decisiontools.org.uk/research/docs/DefiningResearchTable_Oct2017-1.pdf).

### Consent for publication

Not applicable

