## Additional File 1 for "Effectiveness of the BNT162b2 (Pfizer-BioNTech) and the ChAdOx1 nCoV-19 (Oxford-AstraZeneca) vaccines for reducing susceptibility to infection with the Delta variant (B.1.617.2) of SARS-CoV-2"

### S1 Demographics for Population of Interest

Table S1.1: Demographics for population of interest.

| Characteristic | Variable | Summary |
|---|---|---|
| Number of participants | | 2,730,111 |
| Age (in years) | $n$ | 2,730,111 |
| | $n$ missing | 0 |
| | Mean (standard deviation) | 42.1 (23.7) |
| | Median (Q1, Q3) | 42.0 (23.0, 61.0) |
| | Range | 0.0-109.0 |
| Sex | $n$ | 2,730,029 |
| | $n$ missing | 82 |
| | Male, $n$ (%) | 1,360,509 (49.8%) |
| | Female, $n$ (%) | 1,369,520 (50.2%) |
| Ethnicity | $n$ | 2,186,803 |
| | $n$ missing | 543,308 |
| | Black, African, Black British, or Caribbean | 19,674 (0.9%) |
| | Asian or Asian British | 37,353 (1.7%) |
| | Another ethnic group | 83,079 (3.8%) |
| | Mixed or multiple ethnic group | 47,322 (2.2%) |
| | White | 1,999,375 (91.4%) |

Table S1.2: Vaccine doses administered by age group.[1]

| Age Group (years) | $n$ | $n$ missing | No Doses | One Dose | Both Doses |
|---:|---:|---:|---:|---:|---:|
| <18 | 518,696 | 0* | 495,612 (95.5%) | 20,040 (3.9%) | 3,044 (0.6%) |
| 18-39 | 772,847 | 0* | 274,896 (35.6%) | 115,213 (14.9%) | 382,738 (49.5%) |
| 40-59 | 678,147 | 0* | 111,745 (16.4%) | 30,041 (4.4%) | 536,361 (79.1%) |
| 60-79 | 545,019 | 0* | 43,884 (8.1%) | 8,515 (1.6%) | 492,620 (90.4%) |
| ≥80 | 158,964 | 0* | 22,810 (14.3%) | 3,720 (2.3%) | 132,434 (83.3%) |
| All | 2,730,111 | 0* | 954,189 (35.0%) | 178,699 (6.5%) | 1,597,223 (58.5%) |

[1]As extracted on 3rd Sep 2021, not excluding those who died prior to having the vaccine or those who moved into or out of the area over the time period.

*No missing values due to the definition that no recorded values means no doses administered.

### S2 MCMC Parameter Estimates, Trace Plots and Posterior Distributions

Table S2.1: Parameter median estimates and 95% credible interval for all model fits.

| Parameter | Core Model ($\gamma = 1/7$) | Sensitivity to $\gamma$ ($\gamma = 1/3$) | Sensitivity to $\gamma$ ($\gamma = 1/11$) | Double Recovered ($\gamma = 1/7$) |
|---|---|---|---|---|
| $\beta$ | 0.406 [0.3893, 0.4425] | 0.7581 [0.7239, 0.8146] | 0.3104 [0.2953, 0.3448] | 0.4303 [0.4121, 0.4702] |
| $e_{1A}$ | 0.3851 [0.3425, 0.426] | 0.3528 [0.3117, 0.3918] | 0.408 [0.3624, 0.4515] | 0.3637 [0.3194, 0.4058] |
| $e_{2A}$ | 0.6402 [0.614, 0.6647] | 0.6618 [0.6378, 0.6843] | 0.6247 [0.5964, 0.6513] | 0.633 [0.6066, 0.6582] |
| $e_{1P}$ | 0.1954 [0.1041, 0.2809] | 0.263 [0.1815, 0.3399] | 0.1436 [0.0452, 0.2353] | 0.1834 [0.0914, 0.2692] |
| $e_{2P}$ | 0.8392 [0.8212, 0.8559] | 0.8425 [0.8254, 0.8583] | 0.8369 [0.8181, 0.8549] | 0.8365 [0.8183, 0.8534] |
| Initial $I$ | 54.0151 [48.4328, 59.5435] | 46.6017 [40.3953, 52.3173] | 56.2925 [50.9526, 61.6423] | 54.0704 [48.4778, 59.5674] |
| Initial $I_{1A}$ | 14.8932 [10.3012, 19.8382] | 12.8806 [8.042, 18.6471] | 15.6489 [11.369, 20.1298] | 14.8604 [10.2687, 19.7842] |
| Initial $I_{2A}$ | 2.4775 [0.93, 4.7359] | 2.5871 [0.8709, 5.4459] | 2.6046 [1.0844, 4.7457] | 2.4337 [0.9037, 4.739] |
| Initial $I_{1P}$ | 0.7469 [0.1234, 2.0681] | 1.0026 [0.1587, 2.9233] | 0.6512 [0.1119, 1.7435] | 0.7594 [0.1288, 2.1112] |
| Initial $I_{2P}$ | 2.9717 [1.3856, 5.1267] | 3.5131 [1.409, 6.6521] | 2.8499 [1.4196, 4.7717] | 2.9786 [1.3647, 5.1574] |
| $\mu_1$ | 0.8836 [0.4464, 0.9955] | 0.8809 [0.5336, 0.9949] | 0.864 [0.3271, 0.9951] | 0.8809 [0.4259, 0.9957] |
| $\mu_2$ | 0.0545 [0.0021, 0.2734] | 0.0787 [0.0031, 0.3343] | 0.0592 [0.002, 0.309] | 0.0556 [0.0022, 0.2809] |

$\beta$: Transmission Rate, $e_{ij}$ : Effectiveness of $i$ doses of vaccine $j$, Initial $I$: Initial number of unvaccinated infected individuals, Initial $I_{ij}$: Initial number of infected individuals who have received $i$ doses of vaccine $j$, $\mu_i$: Infectiveness of an infected individual with $i$ doses of either vaccine. For number of doses we have $i \in \{1, 2\}$ and for vaccines $j \in \{A, P\}$ for Oxford-AstraZeneca and Pfizer-BioNTech respectively.

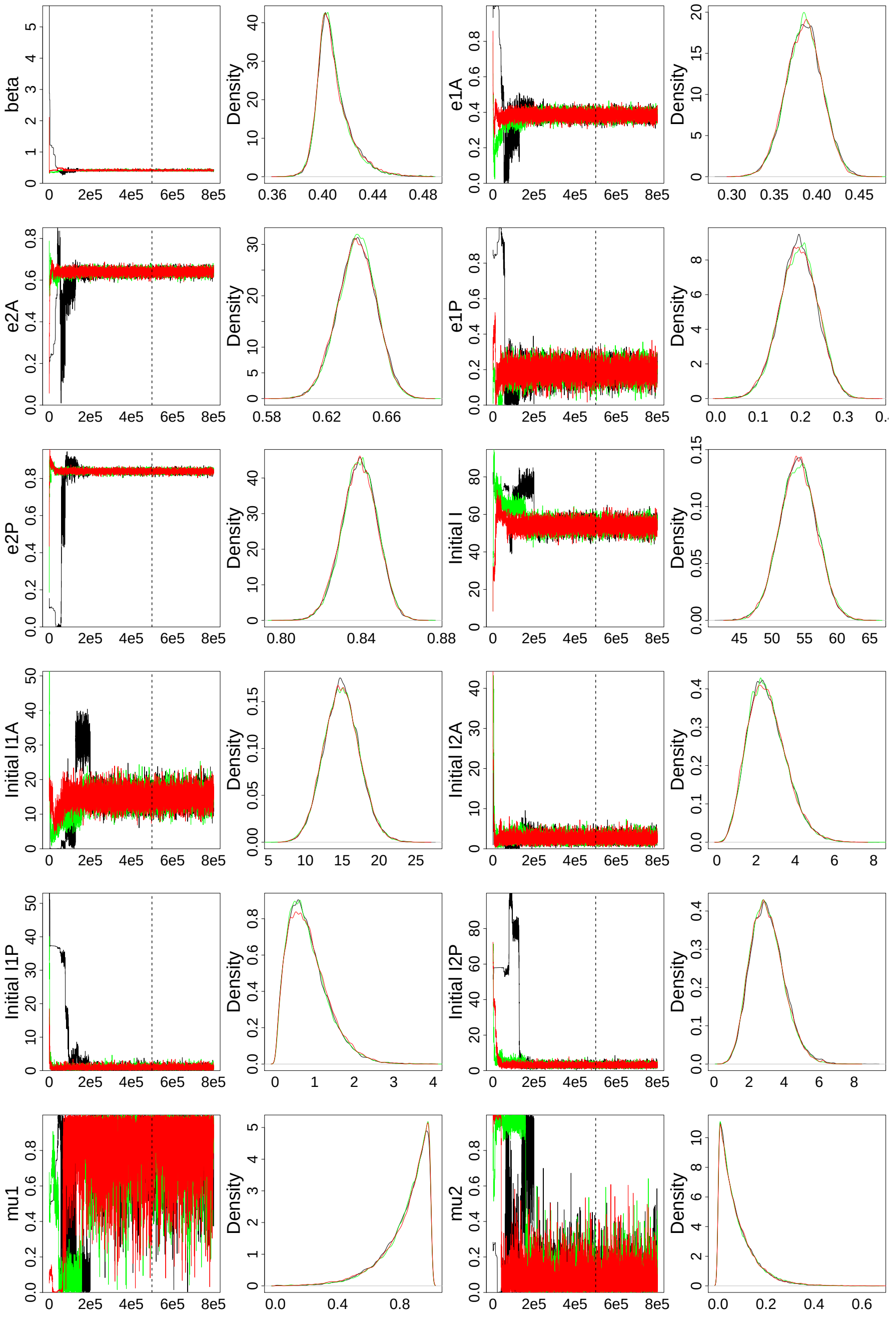


Figure S2.1: MCMC trace and posterior distributions for Core Model fit.

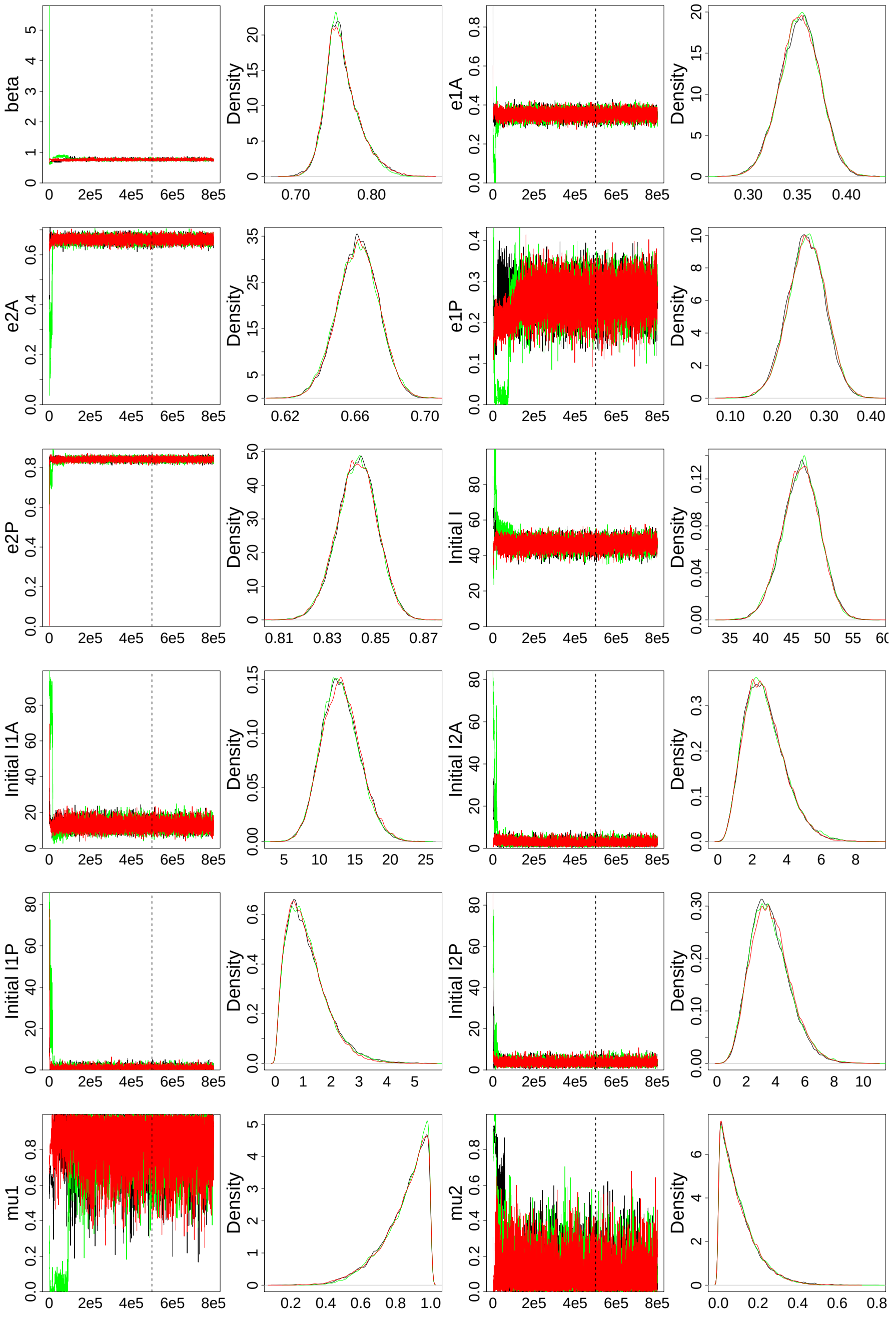


Figure S2.2: MCMC trace and posterior distributions for Sensitivity to $\gamma$ ($\gamma = 1/3$) fit.

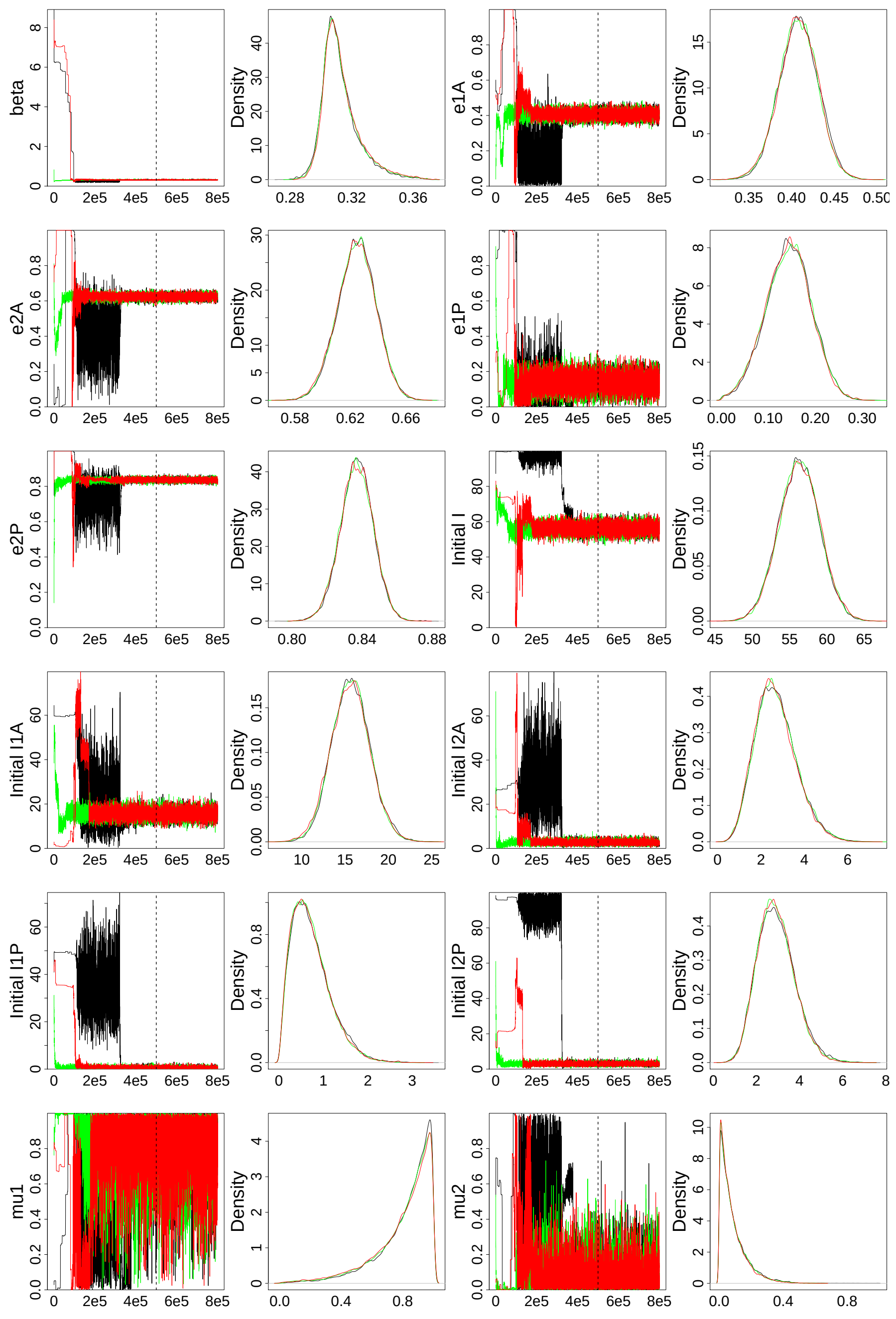


Figure S2.3: MCMC trace and posterior distributions for Sensitivity to $\gamma$ ($\gamma = 1/11$) fit.

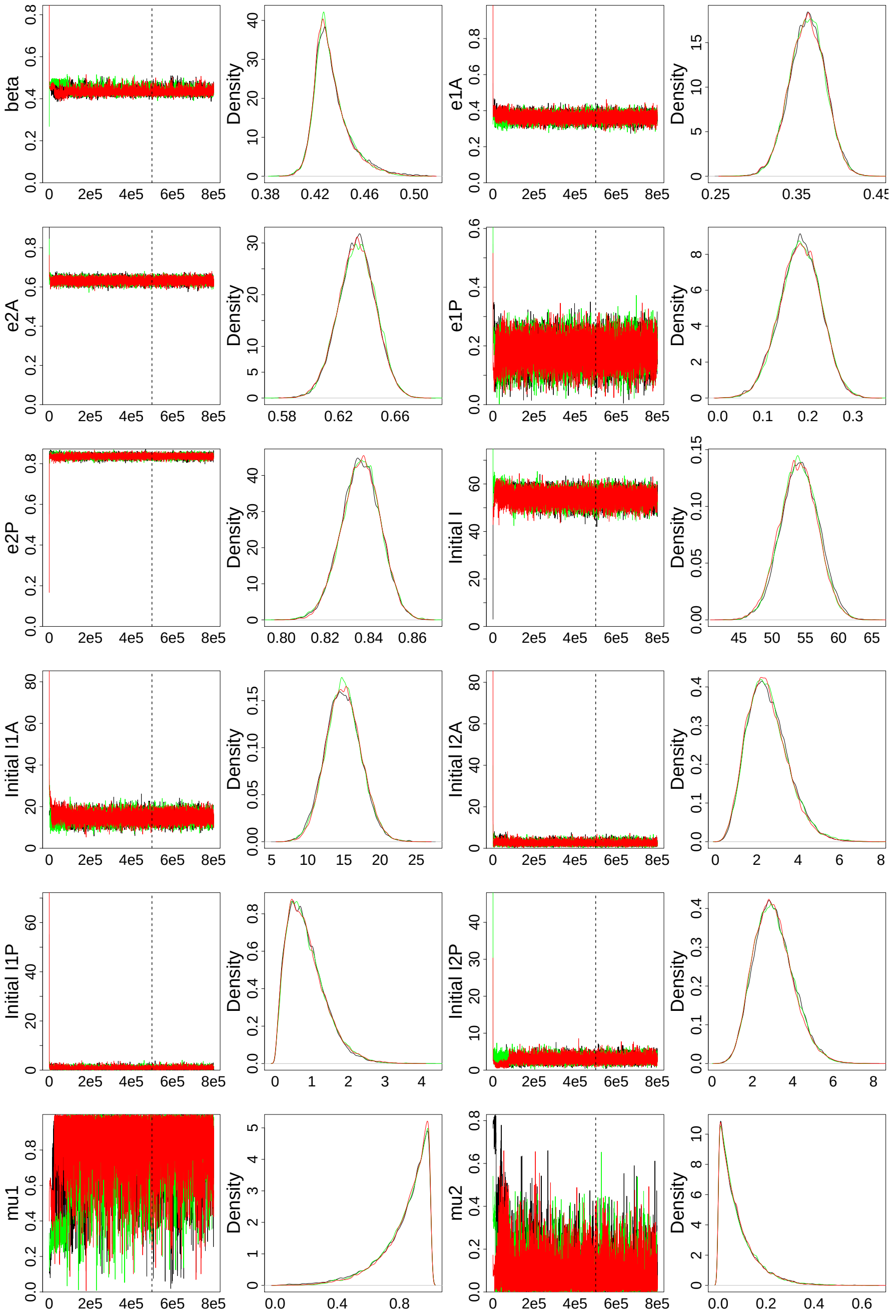


Figure S2.4: MCMC trace and posterior distributions for Double Removed fit.
